# COVID-19 Case Series at UnityPoint Health St. Luke’s Hospital in Cedar Rapids, IA

**DOI:** 10.1101/2020.07.17.20156521

**Authors:** Dianna Edwards, Daniel McGrail

## Abstract

A retrospective, descriptive study of all patients tested for SARS-CoV2 on admission for illness to a community hospital in Iowa from 3/21/20 thru 6/14/20 consisted of evaluation as to demographics, presentation and hospital course. Ninety-one patients were SARS-CoV2 PCR+ with 63% being male and a median age of 60. Cardiovascular disease was a significant comorbidity in the PCR+ group. Fever, cough, dyspnea, nausea, emesis, diarrhea, headache and myalgias were significantly more common in that group, as was an elevated CRP, LDH, serum ferritin and transaminases. Overall survival of the COVID-19 patients was 88%, 77% in the critically ill, 59% of those mechanically ventilated and 33% of those requiring new dialysis. Survival was 93% in those not receiving any antivirals. Survival of those treated with hydroxychloroquine-azithromycin was 92%, compared to 86% of those treated with hydroxychloroquine alone. The latter two groups were significantly more ill than the untreated group. A transition from an early intubation strategy to aggressive utilization of high flow nasal cannula and noninvasive ventilation(i.e BiPAP) was successful in freeing up ICU resources.

## Introduction

The SARS-CoV2 2020 epidemic first became prominent in Iowa in Linn County, where Cedar Rapids is located. On 3/8/2020, the first SARS-CoV2 cases were reported in neighboring Johnson County, which were possibly related to an influx of SARS-CoV2 individuals who had been on an Egyptian cruise^11^. Linn county reported its first case on 3/21/2020 the same day as our first known COVID-19 hospitalized patient at UnityPoint Health(UPH) St. Luke’s Hospital. In this case series, we aim to describe epidemiological, clinical, laboratory, and treatment outcomes of confirmed COVID-19 patients admitted 3/21/2020 thru 6/14/2020 to UPH St. Luke’s Hospital, one of the two hospitals in Cedar Rapids.

## Methods

Using data extracted from the electronic medical record, we performed a retrospective, descriptive study of the first consecutive 91 adults(≥ 17 years) patients admitted to St. Luke’s Hospital with confirmed COVID-19 hospitalized between March 21, 2020(date of first positive case) and June 14, 2020. Cases were confirmed through RT-PCR assays using nasopharyngeal swab specimen collection. Between April 3, 2020 to May 3, 2020 we also collected demographic and patient characteristics on COVID negative adult patients presented to the St. Luke’s Emergency Department with suspected COVID-19 illness. Selection of these patients was based on Emergency Department provider clinical judgement. This comparison data was utilized to help identify risk factors and probability factors for COVID-19 positivity. Outcomes data remains incomplete for patients whom remain hospitalized(n=3). We have excluded patients who died in Emergency Department prior to admission or were admitted to obstetrics with asymptomatic COVID-19 identified on admission screen.

## Results

### Census

Graph 1 demonstrates the daily census at St. Luke’s for hospitalized COVID-19 positive patients and new daily admission. During this study period, we treated 91 patients with the peak census on April 19, 2020 at 21 hospitalized COVID-19 patients. Graph 2 demonstrates ICU resource utilization with peak critical care utilization of ICU and ventilators between April 7^th^ and April 11^th^. This early peak in ICU resources is a reflection of transitioning from an early intubation intervention for COVID-19 related acute hypoxic respiratory failure to a strategy of utilizing high flow nasal cannula and noninvasive ventilation(i.e BiPAP) as supported in the literature^15^.

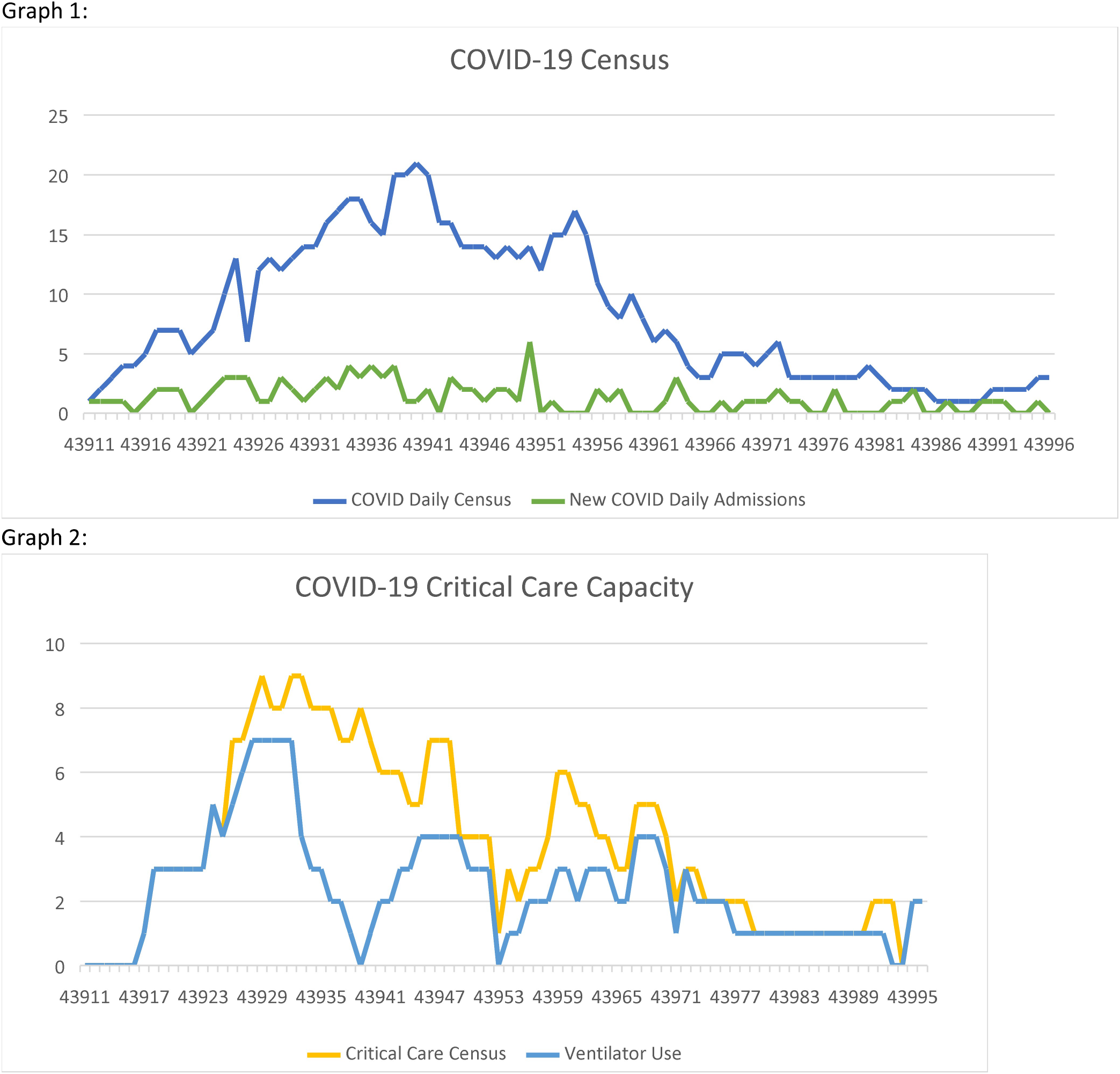

### Demographics and Clinical Characteristics

Table 1 summarizes patient characteristics of COVID positive and negative patients.

**Table 1.** Baseline Demographics and Clinical Characteristics

| Characteristics | No.(%) |  | p |
| --- | --- | --- | --- |
|  | COVID Positive | COVID Negative |  |
| Total | 91(43.5%) | 118(56.5%) |  |
| Sex |  |  |  |
| Male | 57(62.6%) | 71(60.2%) |  |
| Female | 34(37.4%) | 47(39.8%) |  |
| Age, Median, y | 60 | 65 |  |
| Age group, y |  |  |  |
| 17-40 | 18(19.8%) | 8(6.8%) |  |
| 41-60 | 38(41.8%) | 29(24.6%) |  |
| 61-79 | 28(30.8%) | 51(43.2%) |  |
| >80 | 7(7.7%) | 30(25.4%) |  |
| Interpretive Service Use | 24(26.4%) |  |  |
| Employed | 43(47.3%) | 22(18.6%) | 0.001* |
| Group Home | 17(18.7%) | 11(9.3%) | 0.078* |
| Co-morbidities |  |  |  |
| Obesity | 47(51.6%) | 48(40.7%) | 0.150* |
| Hypertension | 47(51.6%) | 77(65.3%) | 0.065* |
| Diabetes Mellitus | 33(36.3%) | 40(33.9%) | 0.834* |
| Cardiovascular Disease | 11(12.1%) | 37(31.4%) | 0.002* |
| Stroke History | 10(11.0%) | 19(16.1%) | 0.391* |
| Smoking in last year | 8(8.8%) | 31(26.3%) | 0.002* |
| Chronic Kidney Disease | 8(8.8%) | 17(14.4%) | 0.305* |
| Chronic Respiratory Disease | 7(7.7%) | 48(40.7%) | 0.001* |
| Immunosuppressed | 3(3.3%) | 7(5.9%) | 0.293** |
| Cancer, Active | 1(1.1%) | 10(8.5%) | 0.015** |
| 1 or more co-morbidities | 49(53.8%) | 97(82.2%) | 0.001* |
| 2 or more co-morbidities | 33(36.3%) | 81(68.6%) | 0.001* |
| Presenting Symptoms |  |  |  |
| Cough | 67(73.6%) | 54(45.8%) | 0.001* |
| Dyspnea | 73(80.2%) | 79(66.9%) | 0.048* |
| Fever | 62(68.1%) | 30(25.4%) | 0.001* |
| Sore Throat | 7(7.7%) | 2(1.7%) | 0.038** |
| Nausea | 32(35.2%) | 17(14.4%) | 0.001* |
| Emesis | 22(24.2%) | 13(11.0%) | 0.019* |
| Diarrhea | 26(28.6%) | 12(10.2%) | 0.001* |
| Headache | 23(25.3%) | 6(5.1%) | 0.001* |
| Myalgias | 40(44.0%) | 10(8.5%) | 0.001* |
| Vitals on Presentation |  |  |  |
| Hypoxia | 69(75.8%) | 86(72.9%) | 0.747* |
| Fever >99.5 | 28(30.8%) | 14(11.9%) | 0.001* |
| Laboratory Findings |  |  |  |
| Lymphopenia(absolute<1300/<br>ul) | 68(74.7%) | 74(62.7%) | 0.090* |
| Thrombocytopenia(<150,000/u<br>l) | 21(23.1%) | 29(24.6%) | 0.930* |
| CRP >6.3 mg/L | 69(75.8%) | 61(51.7%) | 0.001* |
| LDH > 246 U/L | 54(59.3%) | 32(27.1%) | 0.001* |
| Ferritin >388 ng/ml | 48(52.7%) | 19(16.1%) | 0.001* |
| D-Dimer >500 ng/ml | 43(47.3%) | 51(43.2%) | 0.659* |
| D-dimer >1500 ng/ml | 11(12.1%) | 31(26.3%) | 0.018* |
| Procalcitonin> 0.5ng/ml | 13(14.3%) | 21(17.8%) | 0.053* |
| AST or ALT elevation | 50(54.9%) | 34(28.8%) | 0.001* |
| Lactate >2.0 mmol/L | 15(16.5%) | 34(28.8%) | 0.055* |

\*per Chi-Square
\*\*per Fisher's Exact test

During the time period, 91 of the patients tested were positive for COVID-19(44%). Among the 91 patients, 63% were male with a mean age of 61 and median age of 60 years. A large proportion of our admissions were employed(47%, n=43), with 14%(6/43) of those working at food processing plants. Group home residents made up 19% of the COVID-19 patients. Interpretive services (for French, Spanish, Haitian-Creole, Serbian, Nepali, Kurundi, Marchallese, Arabic, and Swahili)were required for 26%(n=24) of the COVID-19 positive patients.

Common co-morbidities among COVID-19 positive patients include obesity(52%), hypertension(52%), and diabetes(36%). A history of stroke(11%), cardiovascular disease(12%), chronic kidney disease(9%), or smoking(9%) were much less common. 46%(n=42) of the COVID-19 patients had no known previous co-morbidity.

The most common presenting symptoms in COVID-19 patients were dyspnea (80%), cough (74%), fever (68%), and nausea(35%). Fever(>99.5°F) was present on admission 31% of the time. We had one patient present with acute embolic stroke, tho after three weeks of cough and fevers. Laboratory findings statistically more commonly present in PCR+ patients included an elevated CRP(76%), an elevated LDH(59%), an elevated serum ferritin(53%) or elevated transaminases(55%). A D-dimer > 1500 ng/ml was actually significantly more common in those patients who tested negative for SARS-CoV2. The majority of those that tested negative presented with symptoms and laboratory abnormalities likely related to their underlying chronic lung disease.

### Hospital Course

Table 2 summarizes the hospital course of the 91 hospitalized COVID-19 positive patients, and outcomes for the 88 discharged.

**Table 2.** Hospital Course and Outcomes of COVID-19 Positive Patients.

|  | No.(%) |  |
| --- | --- | --- |
| Hypoxia at admission | 69(75.8%) |  |
| Required ICU Transfer | 37(40.7%) |  |
| Intubated for Mechanical Ventilation | 19(20.9%) |  |
| Average Days on Vent | 11.35 | Range 1-42 days |
| $\geq 6$L, Vapotherm, BiPAP | 26(28.6%) | |
| New Dialysis required | 9(9.9%) |  |
| Survival |  |  |
| Overall | 77(87.5%) |  |
| Required ICU Transfer | 26(76.5%) |  |
| Intubated for Mechanical Ventilation | 10(58.8%) |  |
| New Dialysis required | 3(33.3%) |  |
| Disposition |  | Average LOS, d |
| Home | 58(65.9%) | 6.08 |
| SNF/Rehab/ICF | 18(20.5%) | 16.7 |
| Deceased | 11(12.5%) | 9.45 |
| Age range, y | 44-91 |  |
| Male | 10(90.9%) |  |

**Table 3.** Treatment and Survival Outcomes for COVID + patients.

|  | No.(%) |  |
| --- | --- | --- |
|  | N (of all discharged) | Survival |
| Untreated | 42(46.2%) | 39(92.9%) |
| Hydroxychloroquine Alone | 21(23.1%) | 18(85.7%) |
| Hydroxychloroquine and Azithro | 12(13.2%) | 11(91.7%) |
| Tocilizumab | 11(12.1%) | 8(80%) |
| Anticoagulation | 45(49.5%) |  |
| High-dose steroids | 31(34.1%) |  |

**Table 4.** Tocilizaumab survival and bacteremia rates in critically ill COVID + patients.

|  | No.(%) |  |  |  |  |  |
| --- | --- | --- | --- | --- | --- | --- |
|  | N | Survival | p | Blood culture Obtained | Bacteremia | p |
| Tocilizumab | 10 | 8(80%) | 0.565* |  | 3(30.0%) | 0.138* |
| No Tocilizumab | 24 | 18(75%) |  |  | 2(8.3%) |  |
| Tocilizumab |  |  |  | 8 | 3(37.5%) | 0.112* |
| No Tocilizumab |  |  |  | 21 | 2(9.5%) |  |

\*per Chi-Square

**Table 5.**
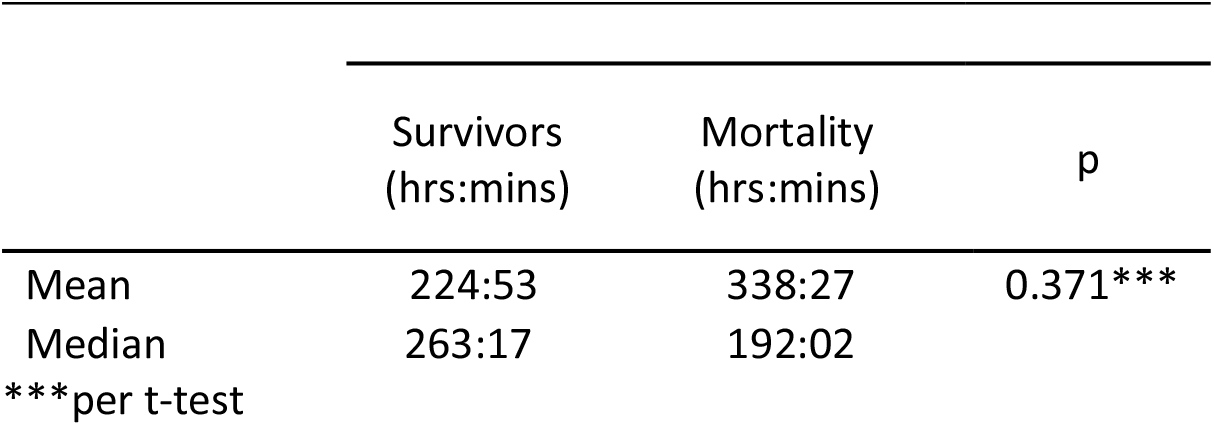
Mechanical ventilation duration and mortality

|  | Survivors<br>(hrs:mins) | Mortality<br>(hrs:mins) | p |
| --- | --- | --- | --- |
| Mean | 224:53 | 338:27 | 0.371*** |
| Median | 263:17 | 192:02 |  |

\*\*\*per t-test

76% of hospitalized patients were hypoxic on admission presentation. 37(41%) of patients required critical care and of those, 89% initially presented with hypoxia. 19(21%) patients developed respiratory failure requiring mechanical ventilation and an additional 26(29%) patients required ≥6L nasal cannula, Vapotherm or BiPAP support. Overall, average days on ventilator was 11.35 days (range 2-42 days). One patient who was ventilated for 42 days, had spent 24 days on a ventilator prior to transfer to St. Luke’s Hospital. Tracheostomy was performed on 2 of the 19 intubated patients. Analysis of the mechanical ventilation duration and survival found that survivors had a lower mean, but a longer median ventilation duration. There was no significant difference in ventilation times between survivors and non-survivors.

Overall survival to hospital discharge was seen in 88%(77/88). 11 patients died, with an age range of 44-91 years old. Ten(90%) of these were males. The critical care survival rate was 78%(26/34). It was 59%(10/17) for those requiring mechanical ventilation. Nine critically ill patients required the initiation of hemodialysis during their hospital stay. Three(33%) survived. Patients discharging to SNF, ICF or inpatient rehab carried the longest average length of stay at 16.7 days.

### 30 Day readmissions

We had 9 readmissions within 30 days of discharge of the 76 discharges. Three patients had 2 separate readmissions, each within 30 days. Ten of the discharged patients have not met their 30 day post discharge milestone.

### Treatment and Survival Outcomes

#### Antivirals

Hydroxychloroquine, azithromycin and convalescent plasma were the antivirals used during this study period. No lopinavir/Kaletra was ordered. Remdesivir was only available for use in one patient who had not been discharged before the end of the study. Early in the epidemic hydroxychloroquine or hydroxychloroquine with azithromycin was used extensively. Patients were counted in the hydroxychloroquine and azithromycin treatment groups if they received a minimum of 3 days of therapy. Usage of hydroxychloroquine and azithromycin dropped off significantly by mid-April with a reliance on supportive, anti-inflammatory, and anticoagulation care. Convalescent plasma was first employed on 4/25/20.

In our study, 46% of hospitalized patients did not receive any antiviral treatment. Those who received hydroxychloroquine alone had a 86% (18/21) survival rate. Those treated with combination hydroxychloroquine and azithromycin had a 92% (11/12) survival rate. The combination(hydroxychloroquine/azithromycin) group was older(median age 58 vs 55 y/o), more commonly hypoxic at admit(100% vs 59%) and had higher Apache 2 scores(mean of 10.3 vs 9.9) than the non-antiviral treated group. The only death in the combination group was in a patient who had been started on those antivirals as an outpatient while she already had a prolonged QT interval. The latter was not monitored during treatment. She died within hours of admission from either a pulmonary embolus or arrhythmia.

In accordance with the Mayo Clinic Expanded Access Program guidelines, data collected on safety and efficacy of convalescent plasma will not be published. This necessitated not reporting 8 patients who received hydroxychloroquine, with or without azithromycin, in addition to their convalescent plasma. By 6/14/20 we had transfused a total of 12 patients with convalescent plasma. In addition, 2 patients received plasma at regional hospitals prior to arrival to St. Luke’s and were included in this review. At the initiation of the convalescent plasma program in April, we had a 4-5 day wait time from order to transfusion of plasma. In May and June as availability of convalescent plasma increased, this wait time was reduced to less than 48 hours.

#### Anti-inflammatories

High dose corticosteroids were used in 34% of all the COVID-19 patients, but in 65% of the ICU patients and in 68% of those requiring mechanical ventilation.

Tocilizumab/Actemra was used in 11(28%) of ICU patients, with 4 receiving a second dose. All 11 patients were critically ill and 8 of the 11 required mechanical ventilation. Eight patients discharged home, 2 died, and 1 remains hospitalized. Secondary infections are a risk with immunomodulators. Both patients that passed away developed bacteremia during their hospital course. There was 1 patient who received Actemra and had evidence of a secondary infection prior to successful home discharge. There was no significant difference in survival between those ICU patients who received tocilizumab/Actemra and those who did not.

#### Anticoagulation

50% of the COVID-19 patients required more than prophylactic anticoagulation doses.

#### Mechanical Ventilation

Nineteen patients required intubation and mechanical ventilation. Of these, two remain in the ICU, on ventilation. Analysis of the required mechanical ventilation durations and survival of the remaining 17, found that survivors had a lower mean, but a longer median ventilation duration. There was no significant difference between survivors and those who perished in ventilation times.

## Summary and Discussion

Our COVID-19 patients demonstrated demographics, risk factors and initial findings similar to those described in the published literature^1,2,6,8,13,14^. As elsewhere noted, we found a moderate male predominance with mean/median ages of 60-61. Hypertension and cardiovascular disease were significant risk factors. Smoking was uncommon. Comorbidities were significantly less common than in the comparison group with 46% of COVID positive patients having no known previous comorbidity at the time of admission. The comparison group, which was selected out on the basis of the emergency department physician’s clinical suspicion, was older with more chronic illnesses, such as chronic respiratory disease. A D-dimer > 1500 ng/ml was actually significantly more common in the comparison group.

The transition away from an early intubation strategy to use of high flow oxygen and noninvasive ventilation resulted in a reduction in critical care and ventilator resource utilization, freeing up limited ICU resources. Through the study period we had an additional 26 patients require ≥6L NC, vapotherm, or BiPAP. Intubation was not required, but would have been recommended in these patients by the prior early intubation guidelines. Although we would have had enough ventilatory equipment to support these patients, other resources such as sedative and paralytic pharmaceutical supplies would have been exhausted. With need for fewer emergent intubations, the dedicated anesthesia COVID intubation team was able to assist and provide support with additional critical care procedures such as proning, central and arterial line placements. In addition, our data indicates a significant portion of the critically ill patients whom were mechanically ventilated required hemodialysis support, 47%(9 of 19). This would predict an additional 12 patients needing dialysis if utilization of high flow nasal cannula or noninvasive ventilation strategies had not been adopted, thus placing a significant amount of strain on resources and potentially exceeding the institutions capacity. Mortality rates for critically ill patients requiring new hemodialysis was high, being >50%.

The mortality rates, while still quite depressingly high, compare well to those reported in the literature^6,10^. The combination of hydroxychloroquine and azithromycin early in the epidemic had a fairly good success rate with few complications. The only death associated with its use had been on unmonitored treatment as an outpatient and died within hours of admission.

Through this study period, knowledge about COVID-19 disease process and investigational treatment strategies were rapidly changing. Examples of pharmacologic and nonpharmacologic changes include utilization of noninvasive ventilation, awake proning strategy, utilization of therapeutic anticoagulation, availability of convalescent plasma and Remdesivir. While these were necessary interventions to maintain most up to date best practice guidelines it does introduce multiple variations in care during this study period.

Our findings also highlight the need for providing a wide range of interpretive services and hospital preparedness to address communication barriers. As noted in the results, 26% of our patients required interpretative services which added a layer of complexity to communicating with patients and their families. Visitor restrictions policy limited family communications to telephone encounters only necessitating a 3 way phone conversation with interpreter services. Virtual interpretive services often could not hear either the patient or the hospital staff through the layers of PPE and noise of HFNC or BiPAP. During a public health crisis, we note the importance of having federal and state public health guidance including hospital discharge instructions available in multiple languages to ensure communication is clear and isolation standards can be followed.

In summary, we hope the knowledge gained through this study will help provide further insights into the epidemiological, clinical, laboratory and treatment outcomes of COVID-19 patients hospitalized in a community-based hospital in Iowa.

## Data Availability

The data that the study is based on is derived from the records of St. Luke's Hospital. The date is in possession of the authors, Dr. McGrail and Dr. Edwards.

